# Hypertension Prevalence and Associated Factors in HIV Positive Women at Kicukiro, Kigali, Rwanda: A Cross-Sectional Analysis

**DOI:** 10.1101/2025.03.13.25323885

**Authors:** Jean Paul Mivumbi, Mojeed Akorede Gbadamosi

## Abstract

**Background:** Hypertension is a significant global health concern, particularly in low- and middle-income countries with limited healthcare access. Women living with HIV (WLHIV) face an increased risk due to prolonged antiretroviral therapy (ART) use, chronic inflammation, and lifestyle factors. This study assessed the prevalence and risk factors of hypertension among WLHIV in Kigali, Rwanda.

**Methods:** A cross-sectional study was conducted among 384 WLHIV aged 18 and older on ART at Kicukiro Health Center. Data were collected using structured questionnaires on demographics, ART use, lifestyle, and health conditions, along with standardized physical measurements. Descriptive statistics and log-binomial regression models were used to determine associations between independent variables and hypertension.

**Results:** The hypertension prevalence was 10.1%. Comorbidities were significantly associated with an increased risk (aPR 6.97, 95% CI 2.93,16.55, *p* < 0.001), as was alcohol consumption (aPR 3.58, 95% CI 1.28,10.07, *p* = 0.015). In contrast, higher education (aPR 0.32, 95% CI 0.13,0.78, *p* = 0.012), contraceptive use (aPR 0.39, 95% CI 0.16,0.91, *p* = 0.030), and work-related physical activity (aPR 0.23, 95% CI 0.06,0.83, *p* = 0.025) were associated with a lower risk.

**Conclusion:** This study suggests the need to integrate hypertension management into HIV care programs, emphasizing comorbidity control, lifestyle modifications, and physical activity promotion. Targeted interventions, such as health education on alcohol risks and cardiovascular health, could help reduce hypertension prevalence among WLHIV in Rwanda, improving long-term health outcomes and quality of life.

## 1. Background

Hypertension is a significant global health concern, particularly in low- and middle-income countries (LMICs), where 82% of the world’s hypertensive population resides (1). It is a major contributor to cardiovascular diseases (CVDs), including heart attacks and strokes, and remains a leading cause of morbidity and mortality worldwide (1). In the context of HIV, advancements in antiretroviral therapy (ART) have successfully extended the life expectancy of people living with HIV (PLHIV). However, this has led to an increased burden of non-communicable diseases (NCDs), including hypertension (2). The co-occurrence of HIV and hypertension presents a significant public health challenge, particularly among women living with HIV (WLHIV), who experience additional risk factors such as hormonal changes, metabolic syndrome, and lifestyle factors (3).

Hypertension, commonly referred to as high blood pressure, is defined as a systolic blood pressure (SBP) of 140 mmHg or higher or a diastolic blood pressure (DBP) of 90 mmHg or higher (1). It results from a combination of modifiable (such as obesity, poor diet, physical inactivity, smoking, alcohol use, and stress) and non-modifiable risk factors (such as genetics, age, and gender) (4). In people living with HIV (PLHIV), additional factors contribute to hypertension, including chronic immune activation, ART-induced metabolic disturbances, and co-infections (5–7). Women living with HIV (WLHIV) are particularly vulnerable, with research indicating higher hypertension prevalence among them compared to HIV-negative women (8,9). Moreover, the interaction between ART and hormonal contraceptives has been suggested as a possible contributor to blood pressure changes (10,11).

Despite advancements in HIV care, hypertension among WLHIV remains underdiagnosed and undertreated, particularly in Sub-Saharan Africa, where the HIV burden is highest(2). Studies report varying hypertension prevalence rates among PLHIV, with 53% in South Africa, 18% in Zambia, and 19.75% across East Africa (12). However, limited research has focused exclusively on WLHIV, and even fewer studies have examined hypertension in this population within Rwanda. Given the high HIV prevalence in Kigali (7%) and the disproportionate burden among women (4% compared to 2% in men), understanding the prevalence and risk factors of hypertension among WLHIV in Rwanda is crucial for developing effective public health interventions(13).

The existing literature highlights several key risk factors associated with hypertension in WLHIV. For instance, long-term ART use has been linked to metabolic syndrome and increased cardiovascular disease risk (10). Lifestyle factors such as excessive alcohol consumption, physical inactivity, and obesity are also significant contributors (4,5). Some studies suggest that contraceptive use may influence blood pressure levels among WLHIV, although findings remain inconsistent (14). Additionally, socioeconomic factors, including education level, income, and healthcare access, play a role in hypertension prevalence (5).

Despite this growing body of evidence, gaps remain in our understanding of hypertension among WLHIV in Rwanda. Furthermore, little is known about the role of ART regimens, contraceptive use, and socioeconomic status in influencing hypertension risk in this population. Addressing these gaps is essential to enhancing integrated HIV and NCD management strategies in Rwanda(15).

This study assessed the prevalence and associated risk factors of hypertension among WLHIV on ART at Kicukiro Health Center in Kigali, Rwanda. By identifying both modifiable and non-modifiable risk factors, the findings will contribute to evidence-based policy recommendations for integrating hypertension management into HIV care programs in Rwanda. Ultimately, the study seeks to support early detection, prevention, and intervention strategies to improve cardiovascular health outcomes among WLHIV.

## 2. Methodology

### Study design, setting, and population

This study employed a cross-sectional research design to assess the prevalence of hypertension among WLHIV receiving ART at Kicukiro Health Center, in Kigali, Rwanda. Data were collected over two months, from March,2 to April,30 2024. Kicukiro Health Center, one of the largest primary healthcare facilities in Kigali. This study enrolled WLHIV aged 18 and older who were receiving HIV care at Kicukiro Health Center. Women under 18, pregnant women, and those with severe cognitive disorders were excluded. The Rwanda national guidelines recommended regular hypertension screening for this high-risk group during medical visits.

### Data source, sample size, and measurements

A structured questionnaire gathered socio-demographic information, ART details, lifestyle factors, and comorbidities. Anthropometric measurements (weight, height, waist, and hip circumferences) and blood pressure were recorded using standardized procedures (14) The sample size was determined using the Kasiulevičius formula for estimating prevalence in cross-sectional studies:

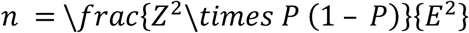

This conservative estimate maximizes the sample size and ensures sufficient statistical power to detect significant associations between hypertension and its risk factors. Given a total WLHIV population of 1,914 at Kicukiro Health Center, the calculated sample size was 384 participants.

To enhance representativeness and generalizability, systematic random sampling was employed, selecting every 5th eligible participant from the facility’s ART patient registry. The first participant was randomly selected from the first five patients arriving each day.

#### Measurement Tools and Procedures

To maintain accuracy and consistency in measurements, this study followed standardized protocols established by the World Health Organization (WHO) and the European Society of Hypertension (ESH). Blood pressure assessments were performed using the AMLON® digital blood pressure monitor, a clinically validated device that meets the validation standards set by both the ESH and the British Hypertension Society (BHS). Measurements were obtained following rigorous procedures to enhance reliability(17). Participants were allowed to rest for a minimum of fifteen minutes before measurement, remaining in a seated position with their arm supported at heart level. Readings were taken from the non-dominant arm, as per standard recommendations. Blood pressure was recorded through three consecutive measurements at five-minute intervals, with the mean of the final two readings utilized for analysis to minimize variability.

Hypertension classification followed WHO guidelines, where normal blood pressure was defined as systolic blood pressure (SBP) below 120 mmHg and diastolic blood pressure (DBP) below 80 mmHg. Pre-hypertension was categorized as SBP ranging from 120 to 139 mmHg or DBP from 80 to 89 mmHg. Hypertension was defined as SBP equal to or exceeding 140 mmHg or DBP equal to or exceeding 90 mmHg.

Anthropometric measurements were conducted in accordance with WHO standardized procedures to ensure accuracy and reliability. Weight was assessed using a calibrated digital scale, height was measured using a stadiometer, and waist and hip circumferences were determined using a non-stretchable measuring tape. Body Mass Index (BMI) was calculated as weight divided by height squared and categorized according to WHO classifications for underweight, normal weight, overweight, and obesity.

Lifestyle factors, including alcohol consumption, smoking history, dietary habits, and physical activity levels, were assessed using a structured questionnaire adapted from WHO’s STEPwise approach to NCD risk factor surveillance. To minimize recall bias, smoking and alcohol consumption data were cross-verified with health records,

Several bias mitigation strategies were implemented to enhance study validity. To reduce selection bias, systematic random sampling was used to ensure an unbiased selection of participants. To address measurement bias, all anthropometric and blood pressure assessments were performed by a single trained nurse following standardized protocols.

#### Study variables definition

The study’s primary outcome was hypertension, characterized by systolic blood pressure readings of 140 mmHg or higher on both days and diastolic blood pressure readings of 90 mmHg or higher. The independent variables included socio-demographic, medical, lifestyle, and anthropometric factors.

Socio-demographic variables consisted of age (categorized as 20–29, 30–39, 40–49, 50–59, and 60 years or older), marital status (single, married, cohabitant, widowed, and divorced), residence (rural or urban), education level (no formal education, primary, secondary/TVET, and tertiary), occupation (unemployed, farmer, business owner, government employee, and others).

Medical Characteristics included women living with HIV on antiretroviral therapy (ART) at Kicukiro Health Center in Kigali, Rwanda, in 2024. ART regimens were classified as ABC/3TC/DTG, AZT/3TC/ATV, TDF/3TC/ATV, TDF/3TC/DTG, and TDF/3TC/EFV. Duration on ART (years) was grouped into 2–6, 7–9, 10–12, 13–15, and 16 years or more. Comorbidities included none, diabetes, HBV, or HCV. Contraceptive use was categorized as Yes or No, and the type of contraceptive used included implant, Intrauterine Device (IUD), injection, oral contraceptive pills, natural methods, and condoms.

Anthropometric Characteristics comprised height (m), weight (kg), waist circumference (cm), hip circumference (cm), waist-to-hip ratio, and body mass index (BMI) (kg/m²).

Lifestyle Characteristics included alcohol intake (never, yes but quit, or yes currently), smoking (never or yes), fruit consumption (I don’t eat fruits or yes), vegetable consumption (I don’t eat vegetables or yes), processed foods consumption (never, sometimes, or always), salt use (never, sometimes, or often), and work-related physical activities (unemployed, yes, or no); Time sitting/reclining in a typical day (minutes) (35,45,60,120, and 180minutes)

### Data analysis

Data from the completed questionnaires were coded in MS Excel and analyzed using SPSS version 27. Descriptive statistics are computed for categorical data, (frequencies and percentages) and continuous data (median and interquartile range). The mean of the last two blood pressure measurements was used to determine hypertension status. A chi-square test was applied to assess the relationship between independent variables and hypertension. Variables with a p<0.2 in bivariate analysis were included in a multivariable Log-Binomial regression to account for confounding. Adjusted prevalence ratios and 95% confidence intervals were provided for each factor of hypertension. A p-value less than 0.05 was considered statistically significant in all analyses.

### Ethical Considerations

Ethical approval was granted by the Mount Kenya University Ethical Review Board with reference MKU/ETHCS/23/01/2024, with additional authorization from Kicukiro Health Center administration. Participants received detailed verbal and written explanations of the study objectives, procedures, and confidentiality measures. For those unable to read, consent forms were read aloud in Kinyarwanda by a trained research assistant, and a thumbprint was obtained in the presence of an independent witness to ensure voluntary participation. Participants were informed of their right to withdraw from the study at any time without any consequences regarding their medical care.

Confidentiality and data protection were maintained throughout the study. Each participant was assigned a unique identifier to replace personal information. All physical records were securely stored, while electronic data were encrypted and password-protected. Data analysis was conducted in a manner that prevented any linkage to individual identifiers, ensuring the anonymity of study participants

## 3. Results

Socio-demographic characteristics of WLHIV on ART out of 385 women living with HIV (WLHIV) included in the sample, Table 1, the median age was 40 years, with the highest proportion (36.4%) aged between 40 and 49 years. Nearly half of the participants (48.1%) were married, and the majority (95.8%) resided in urban areas. Educational attainment varied, with 44.4% completing primary education. Most participants (36.6%) were unemployed.

**Tables 1:** Socio-demographic characteristics of WLHIV on ART at Kicukiro Health center Kigali, Rwanda (n =385)

**Kigali, Rwanda (n =385)**
| **Characteristic** | **Frequency (n)** | **Percentage (%)** |
| --- | --- | --- |
| **Age (years)** |  |  |
| **Medium (IQR)<sup>a</sup>** | 40 (15) |  |
| **Min, Max** | 20, 64 |  |
| 20 - 29 | 81 | 21.0 |
| 30 - 39 | 103 | 26.8 |
| 40 - 49 | 140 | 36.4 |
| 50 - 59 | 51 | 13.2 |
| ≥60 | 10 | 2.6 |
| **Marital status** |  |  |
| Single | 90 | 23.4 |
| Married | 185 | 48.1 |
| Cohabiting | 34 | 8.8 |
| Widowed | 72 | 18.7 |
| Divorced/separated | 4 | 1.0 |
| **Residence** |  |  |
| Rural | 16 | 4.2 |
| Urban | 369 | 95.8 |
| **Education** |  |  |
| No formal education | 122 | 31.7 |
| Primary | 171 | 44.4 |
| Secondary/TVET | 33 | 8.6 |
| Tertiary | 59 | 15.3 |
| **Occupation** |  |  |
| Unemployed | 141 | 36.6 |
| Farming | 54 | 14.0 |
| Business | 47 | 12.2 |
| Government employment | 125 | 32.5 |
| Others | 18 | 4.7 |

### The prevalence of hypertension among WLHIV on ART at Kicukiro Health Center

The prevalence of hypertension among women living with HIV (WLHIV) receiving antiretroviral therapy (ART) at Kicukiro Health Center was **10.1%**

**Figure 1:**
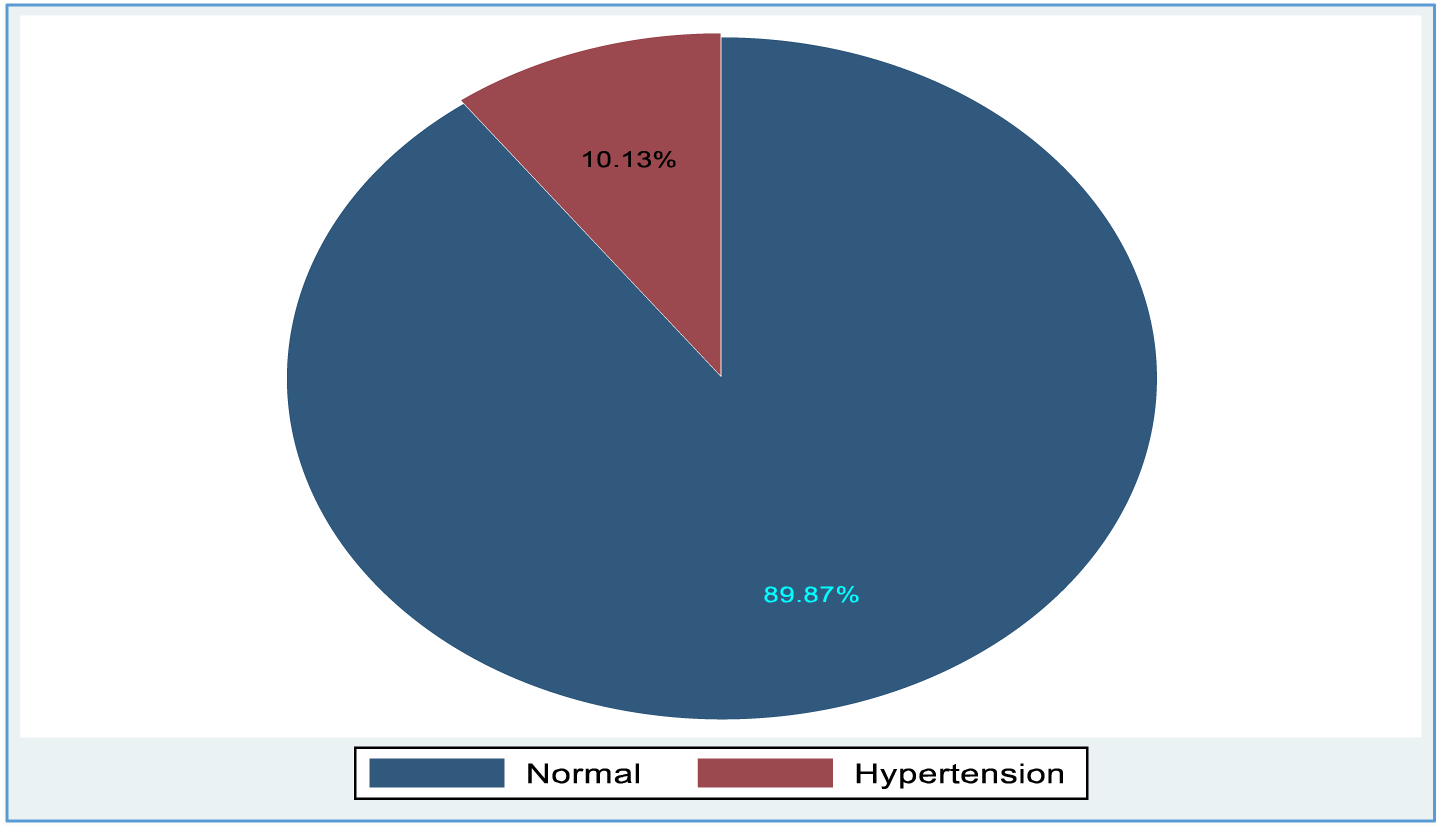
Prevalence of hypertension among women living with HIV on ART

#### Blood Pressure Distribution Among Participants

The systolic and diastolic blood pressure values among the study population are summarized in Table below. The median systolic blood pressure was 125.00 mmHg (IQR: 20.50), ranging from 98.00 to 178.00 mmHg. The median diastolic blood pressure was 76.00 mmHg (IQR: 14.00), with values ranging from 51.00 to 107.00 mmHg

| Characteristics<sup>a</sup> | Mean (SD) | Median (IQR) | Min - Max |
| --- | --- | --- | --- |
| Systolic blood pressure (mmHg) | 127.48 (0.85) | 125.00 (20.50) | 98.00 - 178.00 |
| Diastolic blood pressure (mmHg) | 76.27 (27 (0.55) | 76.00 (14.00) | 51.00 - 107.00 |

IQR, inter-quartile range; Min, Max, minimum, maximum; SD, standard deviation
<sup>a</sup>Variables not normally distributed (Shapiro-Wilk $p < 0.05$ )

### Factors associated with hypertension among WLHIV on ART at Kicukiro Health Center

Bivariate analysis (Table 2) identified several factors significantly associated with hypertension. Among the socio-demographic variables, age and education were notable. Health and lifestyle factors, including alcohol consumption, smoking, processed food intake, salt use, and work-related physical activity, also showed significant associations. Medical factors such as ART regimen, duration of ART use, presence of comorbidities (e.g., diabetes, hepatitis, or other chronic conditions), and contraceptive use were linked to hypertension. Additionally, anthropometric characteristics like body mass index (BMI) and waist-to-hip ratio (WHR) were significantly associated with hypertension.

**Table 2:**
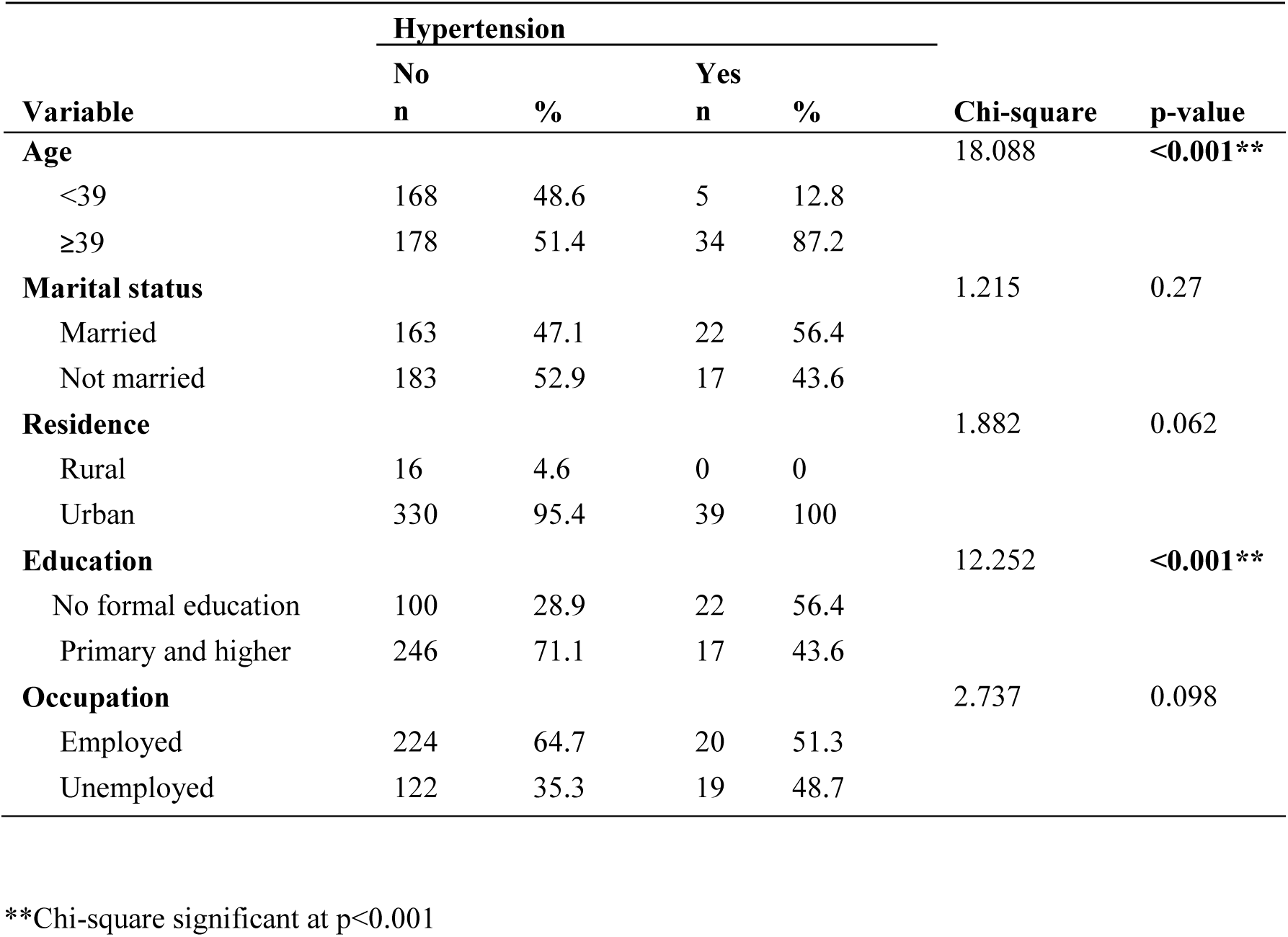
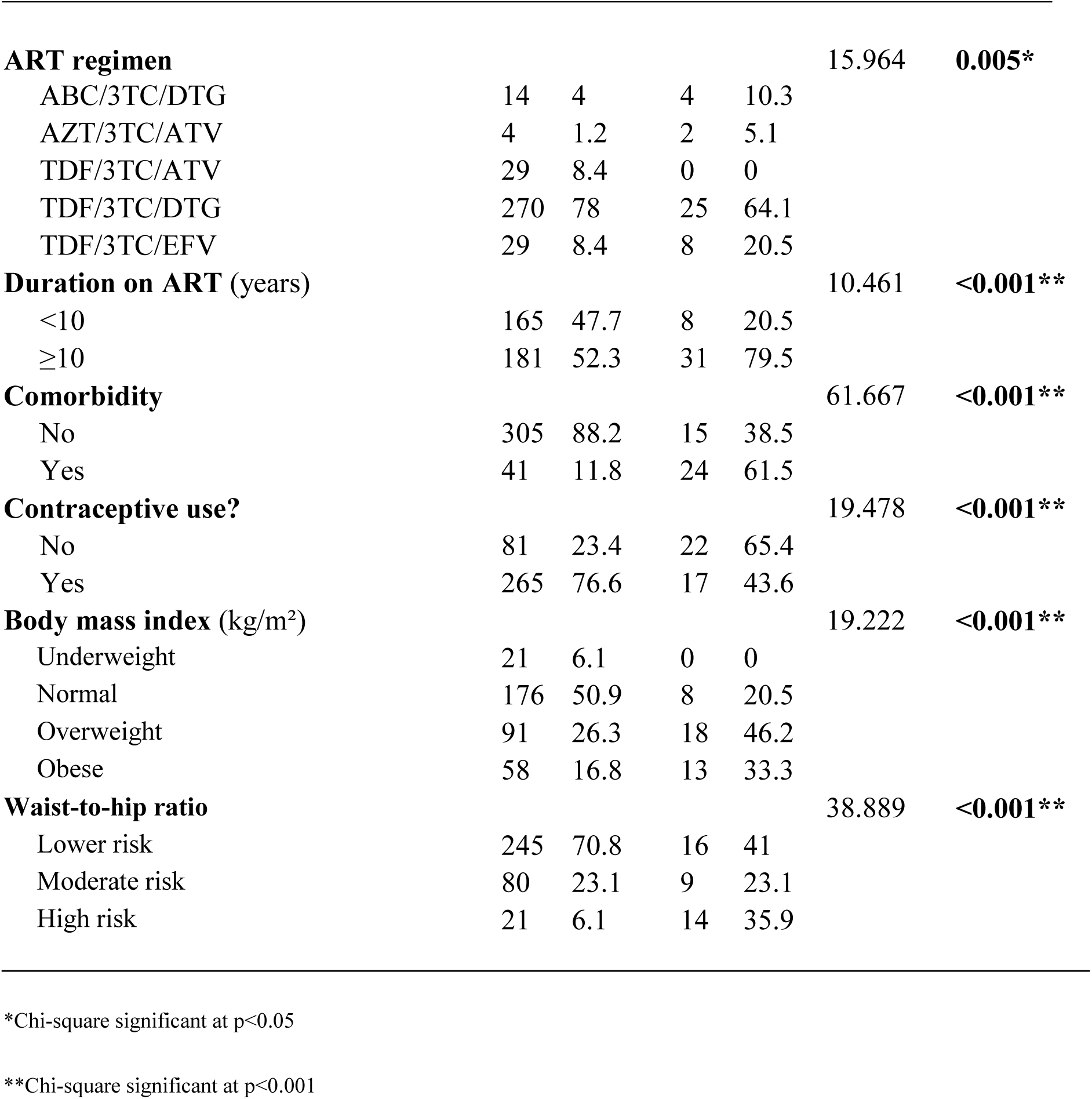
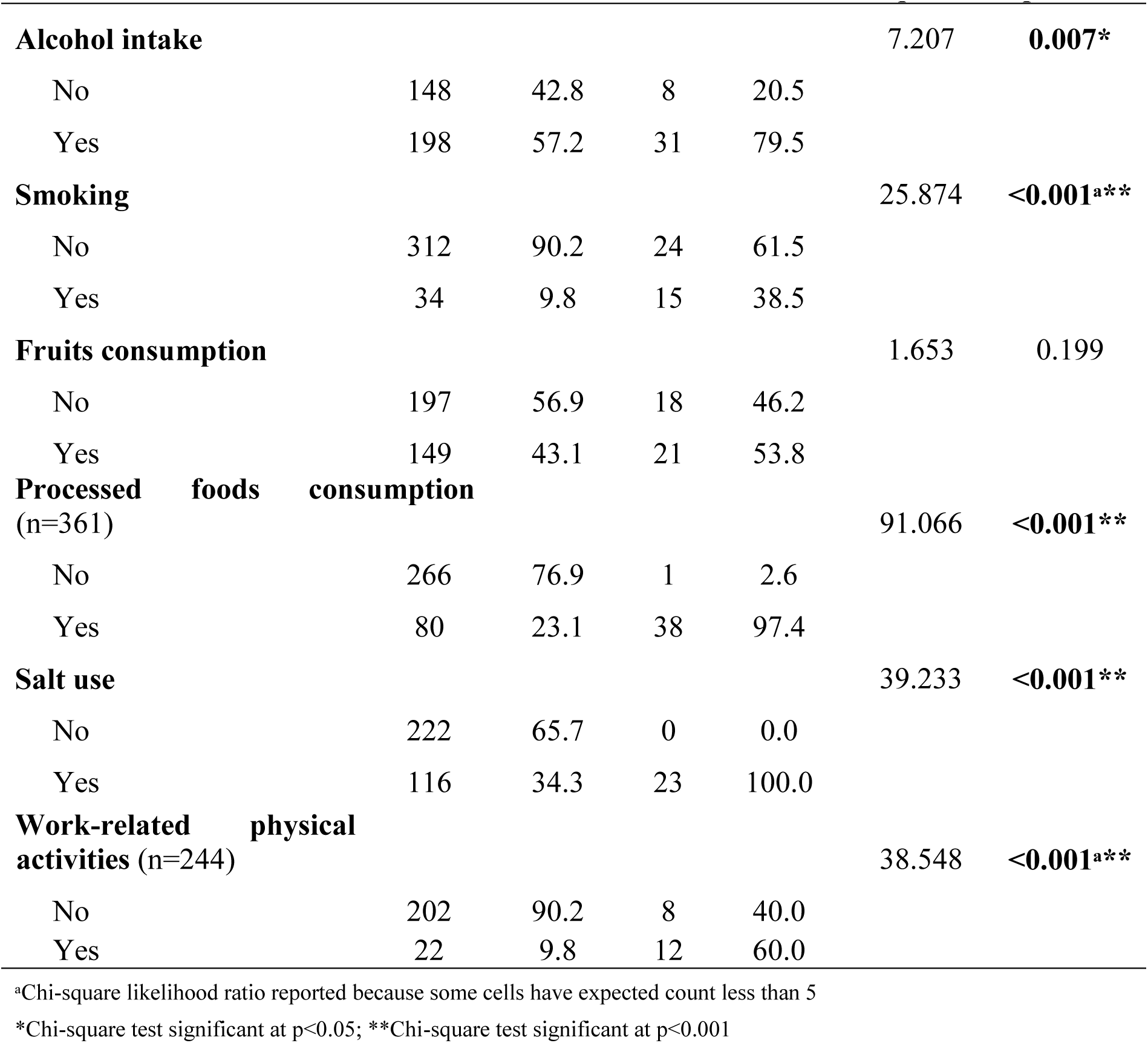
Bivariate association with Hypertension and Socio-Demographic characteristics of WLHIV on ART.

Multivariate analysis (table 3) revealed that women with primary or higher education were less likely to develop hypertension compared to those with no formal education (aPR = 0.32, 95% CI: 0.13 - 0.78, p = 0.012). Regarding health and medical factors, contraceptive use was associated with a lower risk of hypertension compared to non-users (aPR = 0.39, 95% CI: 0.16 - 0.91, p = 0.030). Women with comorbid conditions had a significantly higher likelihood of developing hypertension than those without comorbidities (aPR = 6.97, 95% CI: 2.93 - 16.55, p < 0.001). Among lifestyle factors, alcohol consumption was associated with a higher risk of hypertension compared to non-drinkers (aPR = 3.58, 95% CI: 1.28 - 10.07, p = 0.015). Conversely, engagement in work-related physical activity was linked to a reduced risk of hypertension (aPR = 0.23, 95% CI: 0.06 - 0.83, p = 0.025).

**Table 3:**
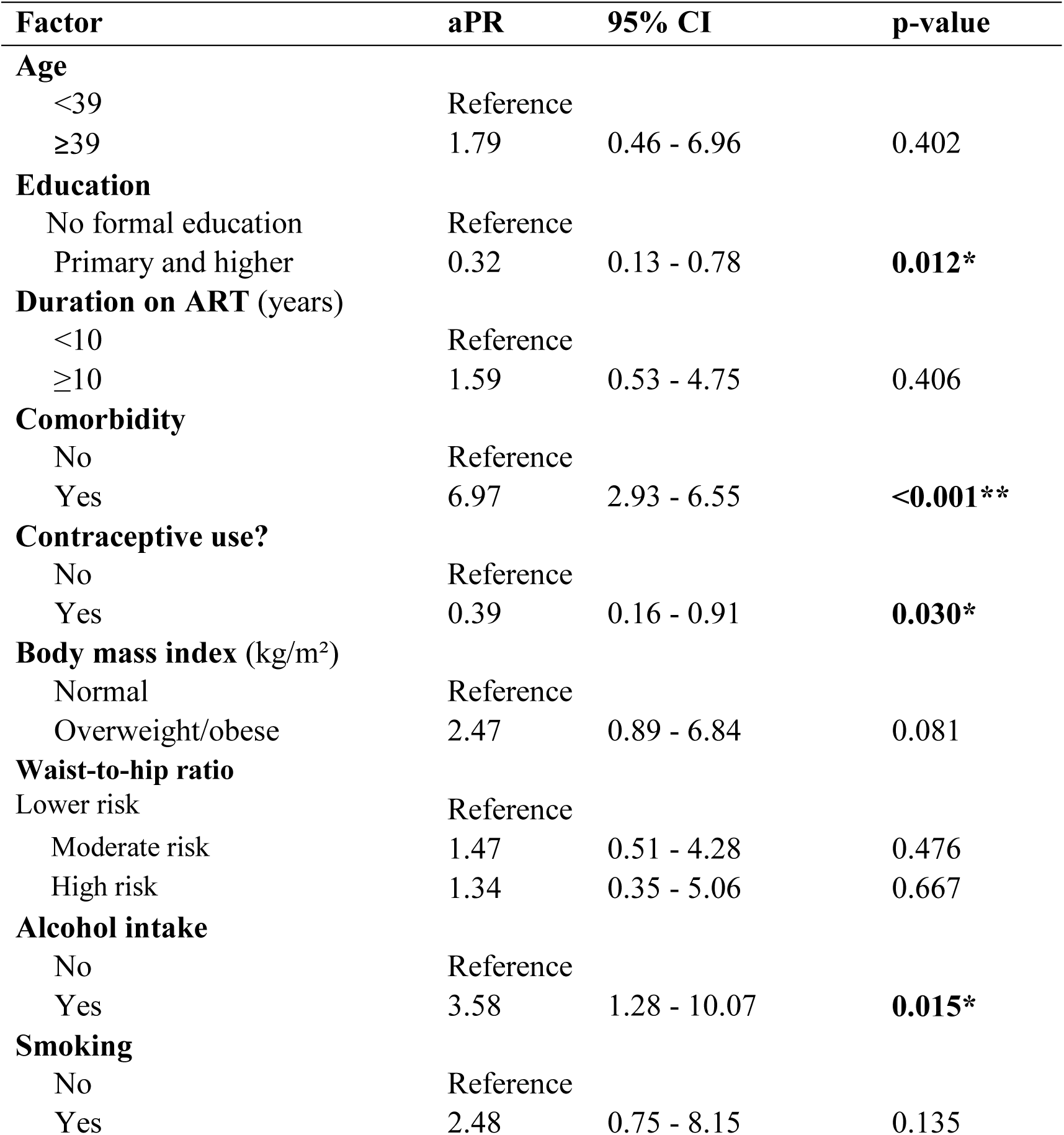
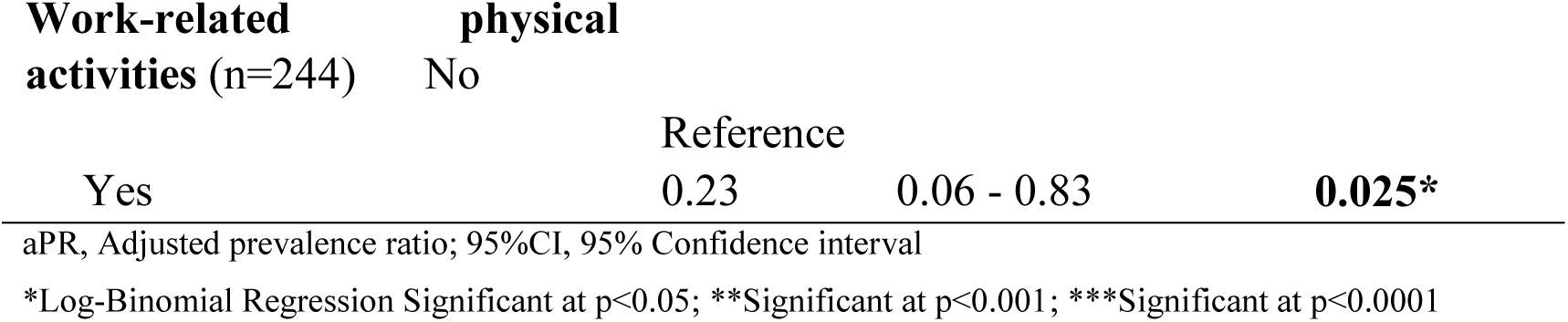
Factors associated with Hypertension among women living with HIV on Antiretroviral Therapy at Kicukiro Health Center, Kigali-Rwanda 2024.

## Discussion

This study examined the prevalence of hypertension and its association with socio-demographic, lifestyle, medical, and anthropometric factors among WLHIV on ART at the Kicukiro Health Center in Kigali, Rwanda, in 2024. The prevalence of hypertension was found to be high among WLHIV and varied according to socio-demographics, Health, and medical factors.

While data on hypertension among WLHIV in Rwanda are limited, the prevalence of 10.13% is notably lower than in other settings within low- and middle-income countries (LMICs). For instance, a study found a prevalence of 23.1% in low-income countries (18). In the region, hypertension rates are significantly higher, with 53% reported in South Africa and 18% in Zambia among women living with HIV (19). A meta-analysis conducted in East Africa reported a prevalence of 19.75% (20), while another southern study indicated a rate of 19.6% in HIV-positive women (21). Additionally, our findings are lower compared to neighboring countries, such as Uganda, where a prevalence of 37% was reported (22), and Burundi, which showed a prevalence of 17.4% among HIV-infected individuals aged 35.4 to 50.2 years (15).

This discrepancy may be attributed to differences in healthcare access and quality over the past two decades. Rwanda has made significant investments in its healthcare system, including widespread access to ART and health education initiatives. Improved healthcare delivery likely contributes to better management of risk factors associated with hypertension compared to countries with less robust healthcare systems. Additionally, the higher proportion of women in our study with primary or higher education may enhance health literacy and awareness of hypertension, leading to more effective preventive measures. The study also indicated a relatively active lifestyle among participants, with over half engaging in work-related physical activities and a low prevalence of smoking (87.3% reported never having smoked). Furthermore, Rwanda’s healthcare system emphasizes the management of HIV and associated comorbidities, such as diabetes and hepatitis, which may help mitigate the prevalence of hypertension. Finally, variations in study design, sample size, and population characteristics can contribute to differences in reported hypertension prevalence across studies.

Our findings highlighted that the education level of WLHIV was associated with hypertension; specifically, individuals with primary or higher education were less likely to develop hypertension. Evidence suggests that differences in educational attainment explain the association between higher health literacy and adherence to lifestyle modifications such as the Dietary Approaches to Stop Hypertension (DASH) diet (23). Additionally, research highlights the importance of social support in combination with health literacy for medication adherence among hypertensive patients. Women with higher educational levels demonstrate greater awareness and preventive behaviors, whereas lower educational attainment is linked to an increased risk of hypertension, higher blood pressure, and greater susceptibility to cardiovascular complications (24).

Furthermore, studies have identified gender differences in the association between education and hypertension, with women showing a stronger correlation between educational level and blood pressure outcomes compared to men (25). Research also suggests that improving health literacy, particularly among women with lower educational backgrounds, may serve as an effective intervention for reducing hypertension-related risks.

Interestingly, in our study, contraceptive use appeared to be protective against hypertension, a finding that contrasts with some studies suggesting hormonal contraceptives may increase blood pressure. This protective effect could be attributed to the more frequent healthcare contact experienced by women using contraceptives. Regular health check-ups may lead to earlier detection and management of elevated blood pressure, thereby preventing the progression of hypertension (26). Moreover, preventive measures such as lifestyle counseling or medication might be initiated earlier in contraceptive users, potentially reducing hypertension risk (26). Additionally, certain progestin-based contraceptives may have a vasodilator effect, helping to regulate blood pressure (27).

Furthermore, HIV status and antiretroviral therapy (ART) use might also influence the relationship between contraceptive use and blood pressure (28). Women on ART who use contraceptives may benefit from more comprehensive medical care, which could mitigate the hypertensive effects often associated with contraceptive use. Therefore, the variation in findings between this study and others may be due to differences in contraceptive types, healthcare access, underlying health conditions, and population characteristics. However, this study did not differentiate between types of contraceptives, and further research is needed to explore hormonal variations and their impact on blood pressure

Our study found that WLHIV with comorbidities were more likely to develop hypertension. Studies from Amsterdam, Netherlands, and Ethiopia similarly show that comorbid conditions such as obesity, diabetes, and dyslipidemia increase the risk of hypertension (29). Long-term use of ART has also been linked to metabolic side effects, including insulin resistance and lipid abnormalities, which further raise this risk (30). Additionally, HIV itself, even when well-managed with ART, can cause chronic immune activation and inflammation, contributing to the development of hypertension. This persistent inflammation leads to vascular damage, making WLHIV more susceptible to cardiovascular diseases (31).

Additionally, our findings suggest that WLHIV who reported alcohol intake were more likely to develop hypertension. Similar studies from Kenya and South Africa have highlighted that alcohol exacerbates liver strain, impairing the body’s ability to regulate blood pressure and manage cholesterol, thus increasing the risk of hypertension. Additionally, alcohol consumption can lead to poor adherence to ART, resulting in suboptimal HIV control, chronic inflammation, and immune activation. Furthermore, alcohol causes vasoconstriction, increasing resistance in blood vessels, which elevates blood pressure (32,33).

Lastly, our findings indicated that work-related physical activity was protective against hypertension. Similar studies conducted in Kenya and South Africa have shown that such activities reduce chronic inflammation, which is particularly beneficial for WLHIV, as it supports immune health (34,35). Regular physical activity also improves insulin sensitivity and helps regulate blood sugar levels, thereby reducing the risk of diabetes, a condition closely associated with hypertension (35,36). Additionally, it aids in controlling lipid levels, further lowering the risk of cardiovascular diseases (36). However, the study did not quantify exercise intensity and duration, making it unclear whether structured physical activity would yield similar benefits. Future research should incorporate objective physical activity measurements, such as accelerometers, to provide more precise data.

## Limitations

This study has several limitations, primarily due to its cross-sectional design, which restricts the ability to establish causal relationships between hypertension and its associated factors. While significant associations were identified, the study cannot determine whether risk factors such as ART duration, alcohol consumption, or comorbidities directly cause hypertension or whether hypertension itself influences these factors. Longitudinal studies are needed to assess the temporal relationship between ART exposure and hypertension development over time.

Additionally, self-reported data on lifestyle behaviors such as alcohol intake, smoking, and physical activity may be subject to recall bias or social desirability bias, as participants might underreport unhealthy behaviors. The study’s urban-based sampling from Kicukiro Health Center may also limit the generalizability of findings to rural populations, where access to healthcare, dietary patterns, and physical activity levels may differ. Future research should include prospective cohort studies and broader geographical sampling to enhance the robustness and applicability of findings.

Despite these limitations, the study provides valuable epidemiological insights into hypertension prevalence among WLHIV in Rwanda, serving as a foundation for future research and policy interventions.

## CONCLUSION

This study reveals a 10.1% prevalence of hypertension among women living with HIV (WLHIV) on antiretroviral therapy (ART) at Kicukiro Health Center in Kigali, Rwanda. Key risk factors identified include comorbidities and alcohol consumption, while higher education, contraceptive use, and work-related physical activity appeared to have a protective effect. These findings underscore the importance of integrating hypertension screening and management into HIV care programs in Rwanda to address the rising burden of non-communicable diseases (NCDs) among WLHIV.

Healthcare providers can enhance HIV care by incorporating routine blood pressure screening and providing lifestyle counseling on alcohol reduction, physical activity, and diet. Integrating HIV and NCD care services would ensure that hypertension management becomes a core component of ART programs, improving patient outcomes. This study aligns with the United Nations Sustainable Development Goals (SDGs), particularly SDG 3 (Good Health and Well-being) and SDG 8 (Decent Work and Economic Growth). It highlights the importance of early detection and integrated care for HIV and hypertension, supporting universal health coverage (SDG 3.8). Addressing hypertension within Rwanda’s HIV care programs improves long-term health outcomes, reduces work-related illness (SDG 8.5, 8.8), and strengthens economic development by promoting a healthier, more productive workforce. Collaboration among healthcare providers, policymakers, and communities is essential to achieving these goals.

## Abbreviations

HIV: Human Immunodeficiency Virus.
WLHIV: Women living with HIV.
ART: Antiretroviral therapy.
NCDs: non-communicable diseases
SDGs: Sustainable Development Goals

## Acknowledgements

Not applicable

## Authors’ contributions

JPM led the drafting of the manuscript and data collections.

MAG data analysis and critical editing for important intellectual content. He also supervised the scientific work throughout the study.

## Availability of data and materials

The datasets generated and analyzed during this study are not publicly available due to confidentiality concerns but can be accessed upon reasonable request from the corresponding author, Jean-Paul Mivumbi, at.

## Declarations

### Ethics approval and consent to participate

This study adhered to ethical guidelines and was approved by the Mount Kenya University Research Ethics Committee (Approval No. MKU/ETHCS/23/01/2024). Permission to conduct the study was also obtained from the administration of Kicukiro Health Center.

All participants provided informed consent before participating in the study. They were thoroughly briefed on the study’s objectives, procedures, and their rights, including confidentiality and the ability to withdraw voluntarily at any stage without facing any consequences. Written consent was obtained from each participant, and all data were managed in strict compliance with ethical standards to ensure privacy and data protection.

## Consent for publication

Not applicable.

## REFERENCE

1. WHO. Hypertension [Internet]. 2023 [cited 2024 Jan 3]. Available from: https://www.who.int/news-room/fact-sheets/detail/hypertension

2. Moyo-Chilufya M, Maluleke K, Kgarosi K, Muyoyeta M, Hongoro C, Musekiwa A. The burden of non-communicable diseases among people living with HIV in Sub-Saharan Africa: a systematic review and meta-analysis. EClinicalMedicine. 2023;65.

3. Abelman RA, Nguyen TTJ, Ma Y, Bacchetti P, Messerlian G, French AL, et al. Body composition changes over the menopausal transition in women with and without Human Immunodeficiency Virus. Clin Infect Dis. 2023;77(2):265–71.

4. Prakash P, Swami Vetha BS, Chakraborty R, Wenegieme TY, Masenga SK, Muthian G, et al. HIV-Associated Hypertension: Risks, Mechanisms, and Knowledge Gaps. Circ Res. 2024;134(11):e150–75.

5. Bigna JJ, Noubiap JJ. Global burden of hypertension in people living with HIV. BMC Med. 2021;19(1):1–2.

6. Derick KI, Khan Z. Prevalence, awareness, treatment, control of hypertension, and availability of hypertension services for patients living with human immunodeficiency virus (HIV) in sub-Saharan Africa (SSA): a systematic review and meta-analysis. Cureus. 2023;15(4).

7. KFF. Global Health Policy. 2023 [cited 2023 Dec 22]. The Global HIV/AIDS Epidemic. Available from: https://www.kff.org/global-health-policy/fact-sheet/the-global-hiv-aids-epidemic/

8. Kasoma Mutebi R, Weil Semulimi A, Mukisa J, Namusobya M, Namirembe JC, Nalugga EA, et al. Prevalence of and factors associated with hypertension among adults on dolutegravir-based antiretroviral therapy in Uganda: a cross sectional study. Integr Blood Press Control. 2023;11–21.

9. Bernard U, Nsanzabera C, Safari E. Prevalence and Associated Factors of Elevated Blood Pressure among People Living with HIV Aged 35-65 Attending Kabutare District Hospital. 2023;

10. Pereira ÍI, Muto AKTM, Dias RFG, de Menezes Filho HR, Fernandes EV, Gouvêa-e-Silva LF, et al. Impact of Metabolic Syndrome and Cardiovascular Risk on the Quality of Life of People Living with HIV. Curr HIV Res. 2024;22(3):170–80.

11. Charkoudian N, Joyner MJ, Johnson CP, Eisenach JH, Dietz NM, Wallin BG. Balance between cardiac output and sympathetic nerve activity in resting humans: role in arterial pressure regulation. J Physiol. 2005;568(1):315–21.

12. Tegegne KD, Adela GA, Kassie GA, Mengstie MA, Seid MA, Zemene MA et al. Prevalence and factors associated with hypertension among peoples living with HIV in East Africa, a systematic review and meta-analysis. BMC Infect Dis. 2023;23(1):724.

13. Hagabimana A, Ndagijimana A, El-Khatib Z, Musafili A, Omolo J, Nzabonimana E, et al. Prevalence of hypertension and associated risk factors among people living with HIV/AIDS in Kigeme, Rwanda 2020. J Interv Epidemiol Public Heal. 2024;7(1).

14. Dzudie A et al. Hypertension among people living with HIV/AIDS in Cameroon: A cross-sectional analysis from Central Africa International Epidemiology Databases to Evaluate AIDS. PLoS One. 2021;22(16):7.

15. Harimenshi D, Niyongabo T, Preux PM, Aboyans V, Desormais I. Hypertension and associated factors in HIV-infected patients receiving antiretroviral treatment in Burundi: a cross-sectional study. Sci Rep. 2022;12(1):20509.

16. Kacanek D, Yee LM, Yao TJ, Lee J, Chadwick EG, Williams PL, et al. Health Outcomes around Pregnancy and Exposure to HIV/Antiretrovirals (HOPE) study protocol: a prospective observational cohort study of reproductive-aged women living with HIV. BMJ Open. 2024;14(7):e084835.

17. Williams B, Mancia G, Spiering W, Agabiti Rosei E, Azizi M, Burnier M, et al. 2018 ESC/ESH Guidelines for the management of arterial hypertension: The Task Force for the management of arterial hypertension of the European Society of Cardiology (ESC) and the European Society of Hypertension (ESH). Eur Heart J. 2018;39(33):3021–104.

18. Zhou B, Carrillo-Larco RM, Danaei G, Riley LM, Paciorek CJ, Stevens GA, et al. Worldwide trends in hypertension prevalence and progress in treatment and control from 1990 to 2019: a pooled analysis of 1201 population-representative studies with 104 million participants. Lancet. 2021;398(10304):957–80.

19. Hines JZ, Prieto JT, Itoh M, Fwoloshi S, Zyambo KD, Sivile S, et al. Hypertension among persons living with HIV—Zambia, 2021; A cross-sectional study of a national electronic health record system. PLOS Glob Public Heal. 2023;3(7):e0001686.

20. Tegegne KD, Adela GA, Kassie GA, Mengstie MA, Seid MA, Zemene MA, et al. Prevalence and factors associated with hypertension among peoples living with HIV in East Africa, a systematic review and meta-analysis. BMC Infect Dis [Internet]. 2023;23(1):724. Available from: 10.1186/s12879-023-08679-x

21. Blair J, Kempf MC, Dionne JA, Causey-Pruitt Z, Wise JM, Jackson EA, et al. Disparities in hypertension prevalence, awareness, treatment, and control among women living with and without HIV in the US South. Open Forum Infect Dis [Internet]. 2023 Dec 18;ofad642. Available from: 10.1093/ofid/ofad642

22. Kanyike AM, Nakawuki AW, Akech GM, Kihumuro RB, Kintu TM, Nalunkuma R, et al. Prevalence, Awareness, and Factors Associated with Hypertension Among People Living with HIV in Eastern Uganda. A Multicentre Cross-Sectional Study. HIV/AIDS - Res Palliat Care [Internet]. 2024 Dec 31;16(null):325–35. Available from: https://www.tandfonline.com/doi/abs/10.2147/HIV.S477809

23. Gusty RP. Health Education Model for Elderly Hypertension on Knowledge, Attitudes, and Adherence to Following the Dietary Approaches to Stop Hypertension (DASH). J Aisyah J Ilmu Kesehat. 2023;8(2).

24. Gaston SA, Forde AT, Green M, Sandler DP, Jackson CL. Racial and ethnic discrimination and hypertension by educational attainment among a cohort of US women. JAMA Netw Open. 2023;6(11):e2344707–e2344707.

25. Ding Y, Wu X, Cao Q, Huang J, Xu X, Jiang Y, et al. Gender disparities in the Association between Educational Attainment and Cardiovascular-kidney-metabolic Syndrome: cross-sectional study. JMIR public Heal Surveill. 2024;10:e57920.

26. Cameron NA, Blyler CA, Bello NA. Oral contraceptive pills and hypertension: a review of current evidence and recommendations. Hypertension. 2023;80(5):924–35.

27. van Lohuizen R, Paungarttner J, Lampl C, MaassenVanDenBrink A, Al-Hassany L. Considerations for hormonal therapy in migraine patients: a critical review of current practice. Expert Rev Neurother. 2024;24(1):55–75.

28. Murray MM, Jensen A, Cieslik T, Cohn SE. Potential risk of drug–drug interactions with hormonal contraceptives and antiretrovirals: prevalence in women living with HIV. Drugs Context. 2020;9.

29. Getahun Z, Azage M, Abuhay T, Abebe F. Comorbidity of HIV, hypertension, and diabetes and associated factors among people receiving antiretroviral therapy in Bahir Dar city, Ethiopia. J comorbidity. 2020;10:2235042X19899319.

30. Ahmadi H, Aghebati-Maleki L, Rashidiani S, Csabai T, Nnaemeka OB, Szekeres-Bartho J. Long-Term Effects of ART on the Health of the Offspring. Int J Mol Sci. 2023;24(17):13564.

31. Abarca YA, Chadalavada B, Ceron JR, Sai BA, Bhatia A, Espinoza I, et al. A Comprehensive Review of the Manifestation of Cardiovascular Diseases in HIV Patients. Cureus. 2025;17(1).

32. Phalane E. A longitudinal investigation of the cardiovascular health of a black South African cohort infected with the human immunodeficiency virus. North-West University (South-Africa); 2021.

33. Syengo SM. LIPID PROFILES, CARDIOVASCULAR DISEASE RISK AND DYSLIPIDEMIA IN HIV-POSITIVE PATIENTS ON HAART AT MACHAKOS LEVEL FIVE HOSPITAL, MACHAKOS COUNTY, KENYA. KENYATTA UNIVERSITY; 2023.

34. Gola M, La Milia D, Cadeddu C, Bardini F, Bianconi B, Bisceglia R, et al. Buffer spaces in healthcare facilities: strategies for managing and designing strategic areas. Popul Med. 2023;5(Supplement):171–2.

35. Garlasco J, Koripalli M, Bridge G. Public health interventions to promote oral health and well-being in patients with type 2 diabetes: a systematic review. Popul Med. 2023;5(Supple).

36. Hayes P, Ferrara A, Keating A, McKnight K, O’REGAN A. Physical activity and hypertension. 2022;

